# No changes in diffusion-weighted imaging-Alberta Stroke Program Early Computed Tomography score from before to after mechanical thrombectomy for anterior circulation occlusion are associated with good outcomes

**DOI:** 10.1101/2022.11.09.22282127

**Authors:** Hiroaki Hashimoto, Tomoyuki Maruo, Yuki Kimoto, Masami Nakamura, Takahiro Fujinaga, Hajime Nakamura, Yukitaka Ushio

**Author notes:** Hiroaki Hashimoto, Neurosurgeon/Medical staff, Department of Neurosurgery, Otemae Hospital, Osaka, 540–0008, Japan, Corresponding Author’s.

## Abstract

**Background:** Although preoperative diffusion-weighted imaging-Alberta Stroke Program Early Computed Tomography Score (DWI-ASPECTS) is well known as a predictor of outcomes after mechanical thrombectomy (MT) for large-vessel occlusion (LVO), assessment of changes in DWI-ASPECT from before to after MT is rare. Therefore, we clarified the relationship between the change in DWI-ASPECTS and clinical outcomes.

**Methods:** In this retrospective single-center study, we enrolled 63 cases of anterior LVOs treated with MT between April 2015 and March 2022. Preoperative and postoperative DWI-ASPECTSs were calculated. DWI-ASPECTSs were categorized into cortical-ASPECTSs (c-ASPECTSs) and subcortical ASPECTSs and assessed. Additionally, medical variables related to patients, such as sex, age, National Institutes of Health Stroke Scale (NIHSS) score, and premorbid modified Rankin Scale (mRS) score, were evaluated. A good outcome was defined as an mRS score of 0 or 2 at 3 months.

**Results:** Forty-five patients met the inclusion criteria. Among the patients, 9 (20%) had a good outcome. The good outcome group showed significantly higher postoperative DWI-ASPECTs (median 8 vs. 5, *p* = 0.012) and c-ASPECTSs (median 4 vs. 3, *p* = 0.020) than the not-good outcome group. No changes in DWI-ASPECTSs and c-ASPECTSs from before to after MT were significantly associated with the good outcome (*p* = 0.017, *p* = 0.016, respectively). The cut-off values for the good outcome on receiver operating characteristic curve analysis for differences between DWI-ASPECTSs and c-ASPECTSs was 0 [area under the curve (AUC) 0.77, sensitivity 0.67, specificity 0.78] and 0 [AUC 0.74, sensitivity 0.44, specificity 1.00]. Logistic regression analyses showed that baseline NIHSS score (odds ratio, 0.69; 95% confidence interval 0.48–1.00; *p* = 0.046) and postoperative DWI-ASPECTS (odds ratio, 2.27; 95% confidence interval 1.02–5.04; *p* = 0.039) were independent factors for the good outcome.

**Conclusions:** The good outcome of patients with anterior LVO was not associated with any changes in DWI-ASPECTSs and c-ASPECTSs from before to after MT.

## Introduction

Diffusion-weighted imaging (DWI)-Alberta Stroke Program Early Computed Tomography Score (ASPECTS) and DWI lesion volume (Vol_DWI_) are negatively correlated ^1,2^, and in acute ischemic stroke (AIS) with large-vessel occlusion (LVO), DWI-ASPECTS 4 and Vol_DWI_ 71 mL were suggested as cut-off values for the prediction of a poor outcome ^3^. Previous studies reported that mechanical thrombectomy (MT) was effective for LVO patients who were within 6–12 hours from the onset and showed ASPECTS ≥6 or 7/DWI-ASPECTS ≥6 ^4-8^, and recently, DAWN ^9^ and DEFUSE 3 ^10^ trials suggested that even in LVO over 6 hours from the onset, MT was effective for patients whose ischemic core volume was less than 51 mL or 70 mL, respectively. Therefore, tissue-based evaluation of AIS is indispensable for the optimal indication of MT, and the correlation between ASPECTS and ischemic core volume was fair ^11^. Therefore, high ASPECTS/DWI-ASPECTS is reported to be a predictor of good outcomes among patients undergoing MT ^5,7,12^. In the American Heart Association (AHA)/American Stroke Association (ASA) guideline, MT is recommended for AIS-LVO based hours of symptom onset and ASPECTS, and MT is not recommended in AIS-LVO with ASPECTS <6 ^13^. At our institution, MT has been chosen according to the guidelines, expecting good results. Unfortunately, although we adhered to these guidelines, some AIS patients undergoing MT showed poor outcomes. Additionally, we have frequently experienced DWI-ASPECTS worsening from before to after MT. Therefore, many studies focused on DWI-ASPECTS before MT to reveal factors to predict good or poor outcomes and to help physicians select AIS-LVO patients who would benefit from MT ^11,14-20^. However, studies investigating DWI-ASPECTS after MT are few, and it remains unclear how changing DWI-ASPECTS from before to after MT relates to the outcomes of AIS patients undergoing MT.

In this study, we enrolled patients with anterior circulation occlusion-related AIS who had MT treatment to clarify factors related to good outcomes after MT by investigating changes in DWI-ASPECTSs from before to after MT. Prediction of outcomes after MT is crucial for physicians to plan the treatment and rehabilitation for AIS patients.

## Methods

### Patients and study setting

In this retrospective study, we enrolled patients who underwent MT because of AIS at Otemae Hospital between April 2015 and March 2022. We referred to the AHA/ASA guideline ^13^, and the following criteria for patient enrollment were set: (1) underwent magnetic resonance imaging (MRI) both before and after MT and completion of postoperative MRI within 21 postoperative days; (2) occlusion of anterior circulation including common carotid artery (CCA), internal carotid artery (ICA), M1 and M2 segments of the middle cerebral artery (MCA); (3) MT within 24 hours of AIS onset; (4) preoperative DWI-ASPECTS ≥6 or preoperative DWI-ASPECTS <6 within 6 hours of AIS onset; (5) patients with clinical outcomes obtained at 3 months after onset. If the symptom onset was unknown, the onset was defined as the time when patients were last known to be in wellness. The Ethics Committee of the Otemae Hospital (Osaka, Japan, approval no. CT210421002) gave the ethical approval for this work, and it was conducted following the Declaration of Helsinki guidelines for experiments involving humans. Informed consent was obtained using the opt-out method from our center’s website because of the retrospective and noninvasive nature of the study.

### Data collection

We retrospectively collected and evaluated medical variables related to patients [sex, age, National Institutes of Health Stroke Scale (NIHSS) score ^21^, and premorbid modified Rankin Scale (mRS) score ^22^]. Imaging findings (laterality of AIS and arterial occlusion site), treatment details [retrieval attempts, onset to reperfusion time, modified Thrombolysis in Cerebral Infarction (mTICI) scale grade, and combination with tissue plasminogen activator (tPA)], and outcomes [mRS score three months after onset, hemorrhagic infarction, and subarachnoid hemorrhage (SAH)] were included. We defined a good outcome as an mRS score 0 to 2 at 3 months ^23^ and successful reperfusion after MT as mTICI grade 2b or 3 ^24^. Two stroke specialists certified by the Japan Stroke Society independently evaluated DWI-ASPECTS (H.H. and T.M.). The DWI-ASPECTS was categorized into subcortical ASPECTS (sc-ASPECTS) with five structures (the caudate, lentiform nucleus, internal capsule, insular ribbon, and white matter) and cortical ASPECTS (c-ASPECTS) with six structures in MCA cortical regions^23^. For quantitative assessment of Vol_DWI_, Digital Imaging and Communications in Medicine (DICOM) DWI-MRI date were imported to MATLAB R2020b (MathWorks, Natick, MA, USA), and lesions identified by DWI were segmented manually using the image segmenter app in MATLAB (https://www.mathworks.com/help/images/ref/imagesegmenter-app.html). These procedures enabled us to calculate Vol_DWI_.

### Statistical analyses

Categorical data and continuous variables are presented as patients (percentages) or median (interquartile range (IQR)), respectively. Clinical differences between the good and not-good outcome groups were assessed using the chi-squared test for categorical variables. The two-tailed Wilcoxon rank-sum test was used for continuous variables. The correlation coefficient was calculated using Spearman correlation analysis. The receiver operating characteristic (ROC) curve was used to perform the sensitivity and specificity. The cut-off value (COV) was defined as the maximal Youden index (sensitivity + specificity – 1). A logistic regression model was used to calculate the odds ratios (OR) with 95% confidence intervals (CIs). The significance level was 0.05. All statistical analyses were performed using the Statistical and Machine Learning Toolbox of MATLAB R2020b (MathWorks, Natick, MA, USA).

### Data availability

All data in this study are available from the corresponding authors upon reasonable request and after additional ethics approval.

## Results

### Baseline characteristics

Forty-five patients fulfilled the inclusion criteria of the 63 patients treated within the study period. The median (IQR) age was 76 years (62–86), and 47% were females. The frequency of preoperative DWI-ASPECTS < 6 was 7 (16%). The rate of good clinical outcomes was 20% (9/45 patients). Successful reperfusion, defined as mTICI grade 2b or 3, was achieved in 82%. Table 1 indicates the baseline characteristics, procedural variables, and complications between the good and not-good outcome groups. No significant differences were noted between the two groups regarding the sex distribution, premorbid mRS score, laterality, arterial occlusion site, retrieval attempts, onset to reperfusion time, mTICI grade, combination with tPA therapy, frequency of preoperative DWI-ASPECTS < 6, and incidence of SAH. However, in the good outcome group, the age and NIHSS scores were significantly lower than those in the not-good outcome group (*p* = 0.00097 and 0.00016, respectively: Wilcoxon rank-sum test). Additionally, the percentage of good outcomes in hemorrhagic infarctions was statistically lower than that in no hemorrhagic infarctions (8% vs. 33%, *p* = 0.036: chi-squared test).

**Table. 1.** Descriptive statistics regarding baseline, procedural, and outcome parameters.

| Characteristics | Patients n (%) | Good outcome<br>(% or IQR) | Not-Good outcome (%<br>or IQR) | *p*-value |
| --- | --- | --- | --- | --- |
| **Sex** |  |  |  |  |
| Male | 24 (53) | 6 (25) | 18 (75) | 0.37 |
| Female | 21 (47) | 3 (14) | 18 (86) |  |
| **Age (years)** |  | 61 (49 – 71) | 83 (76 – 89) | *0.00097 |
| **NIHSS** |  | 12 (6 – 14) | 19 (17 – 22) | *0.00016 |
| **Premorbid mRS score** |  | 0 (0 – 1) | 1 (0 – 3) | 0.18 |
| **Laterality** |  |  |  |  |
| Left | 25 (56) | 4 (16) | 21 (84) | 0.45 |
| Right | 20 (44) | 5 (25) | 15 (75) |  |
| **Arterial occlusion site** |  |  |  |  |
| CCA + ICA + MCA-M1 | 32 (71) | 8 (25) | 24 (75) | 0.19 |
| MCA-M2 | 13 (29) | 1 (8) | 12 (92) |  |
| **Retrieval attempts** |  | 1 (1-3) | 2 (1-2) | 0.98 |
| **Onset to reperfusion time (hours)** |  | 8 (4-11) | 5 (4-8) | 0.25 |
| **mTICI grade** |  |  |  |  |
| 2b + 3 | 37 (82) | 7 (19) | 30 (81) | 0.70 |
| Others | 8 (18) | 2 (25) | 6 (75) |  |
| **tPA therapy** |  |  |  |  |
| Yes | 18 (40) | 2 (11) | 16 (89) | 0.22 |
| No | 27 (60) | 7 (26) | 20 (74) |  |
| **Preoperative DWI-ASPECTS** |  |  |  |  |
| ≥6 | 38 (84) | 9 (24) | 29 (76) | 0.15 |
| <6 | 7 (16) | 0 (0) | 7 (100) |  |
| **Hemorrhagic infarction** |  |  |  |  |
| Yes | 24 (53) | 2 (8) | 22 (92) | *0.036 |
| No | 21 (47) | 7 (33) | 14 (67) |  |
| **SAH** |  |  |  |  |
| Yes | 13 (29) | 1 (8) | 12 (92) | 0.19 |
| No | 32 (71) | 8 (25) | 24 (75) |  |

Patients (%) or median (IQR) are demonstrated for categorical data or continuous variables, respectively. *P* values were calculated using the chi-squared test for categorical variables and the Wilcoxon rank-sum test for continuous variables. Statistically significant *p* values are flagged with an asterisk.
ASPECTS indicates Alberta Stroke Program Early Computed Tomography Score; CCA, common carotid artery; DWI, diffusion-weighted imaging; ICA, internal carotid artery; IQR, interquartile range; MCA-M1, middle cerebral artery-M1 segment; MCA-M2, middle cerebral artery-M2 segment; mRS, modified Rankin Scale; mTICI, modified Thrombolysis in Cerebral Infarction; NIHSS, National Institutes of Health Stroke Scale; SAH, subarachnoid hemorrhage; and tPA, tissue plasminogen activator.

### DWI-ASPECTS and Vol_DWI_

There were significant negative correlations between DWI-ASPECTS and Vol_DWI_ before (correlation coefficients; −0.88, *p* = 1.8 × 10^−15^) and after MT (correlation coefficients; −0.86, *p* = 2.6 × 10^−14^). Figure 1 indicates the box-and-whisker plot between DWI-ASPECTS and Vol_DWI_ before and after MT. In the preoperation and postoperation, all Vol_DWI_ of DWI-ASPECTS ≥7 was less than 70 mL. Preoperative DWI-ASPECTS ≥6 showed uneven distribution; however, in postoperation, DWI-ASPECTS <6 increased. In preoperation, DWI-ASPECTS, c-ASPECTS, and Vol_DWI_ values showed no significant differences between the good and not-good outcome groups (Table 2). However, postoperative DWI-ASPECTS and c-ASPECTS were significantly higher in the good outcome group than in the not-good outcome group (*p* = 0.012 and 0.020, respectively: Wilcoxon rank-sum test), and postoperative Vol_DWI_ was significantly lower in the good outcome group than in the not-good outcome group (*p* = 0.0057, Wilcoxon rank-sum test).

**Table. 2.** Pre- and postoperative DWI-ASPECTS

| Characteristics | Patients n (%) | Good outcome (%) | Not-good outcome (%) | *p*-value |
| --- | --- | --- | --- | --- |
| **Days of postoperative MRI (days)** |  | 1 (1 – 6) | 1 (1 – 5) | 0.95 |
| **Preoperation** |  |  |  |  |
| **DWI-ASPECTS** |  | 8 (7 – 9) | 7 (6 – 9) | 0.47 |
| **sc-ASPECTS** |  | 4 (3 – 4) | 3 (2 – 4) | 0.64 |
| **c-ASPECTS** |  | 4 (4 – 6) | 5 (2 – 6) | 0.56 |
| **Vol<sub>DWI</sub> (mL)** |  | 9 (6.3 – 35.0) | 27 (4.6 – 59.4) | 0.31 |
| **Postoperation** |  |  |  |  |
| **DWI-ASPECTS** |  | 8 (7 – 9) | 5 (3 – 8) | *0.012 |
| **sc-ASPECTS** |  | 3 (3 – 4) | 2 (1 – 4) | 0.097 |
| **c-ASPECTS** |  | 4 (4 – 6) | 3 (0 – 4) | *0.020 |
| **Vol<sub>DWI</sub> (mL)** |  | 13 (6.8 – 23.4) | 57 (21.8 – 123.2) | *0.0057 |
| **Differences** |  |  |  |  |
| **Pre - Postoperative DWI-ASPECTS** |  | 0 (0 – 0) | 1 (0 – 3) | *0.011 |
| **No changes (differences = 0)** | 16 (36) | 6 (37.5) | 10 (62.5) | *0.017 |
| **Worsening (differences > 0)** | 26 (58) | 2 (8) | 24 (92) |  |
| **Pre - Postoperative sc-ASPECTS** |  | 0 (0 – 0) | 0 (0 – 1) | 0.20 |
| **Pre - Postoperative c-ASPECTS** |  | 0 (0 – 0) | 0 (0 – 2) | *0.013 |
| **No changes (differences = 0)** | 27 (60) | 8 (30) | 19 (70) | *0.016 |
| **Worsening (differences > 0)** | 16 (36) | 0 (0) | 16 (100) |  |
| **Post - Preoperative Vol<sub>DWI</sub> (mL)** |  | 1 (-2.7 – 3.1) | 18 (4.8 – 55.0) | *0.00086 |

Patients (%) or median (IQR) are demonstrated for categorical data or continuous variables, respectively. *P* values were calculated using the chi-squared test for categorical variables and the Wilcoxon rank-sum test for continuous variables. Statistically significant *p* values are flagged with an asterisk.
ASPECTS indicates Alberta Stroke Program Early Computed Tomography Score; c-ASPECTS, cortical ASPECTS; DWI, diffusion-weighted imaging; IQR, interquartile range; MRI, magnetic resonance imaging; sc-ASPECTS, subcortical ASPECTS; and Vol<sub>DWI</sub>, DWI lesion volume.

**Fig. 1.**
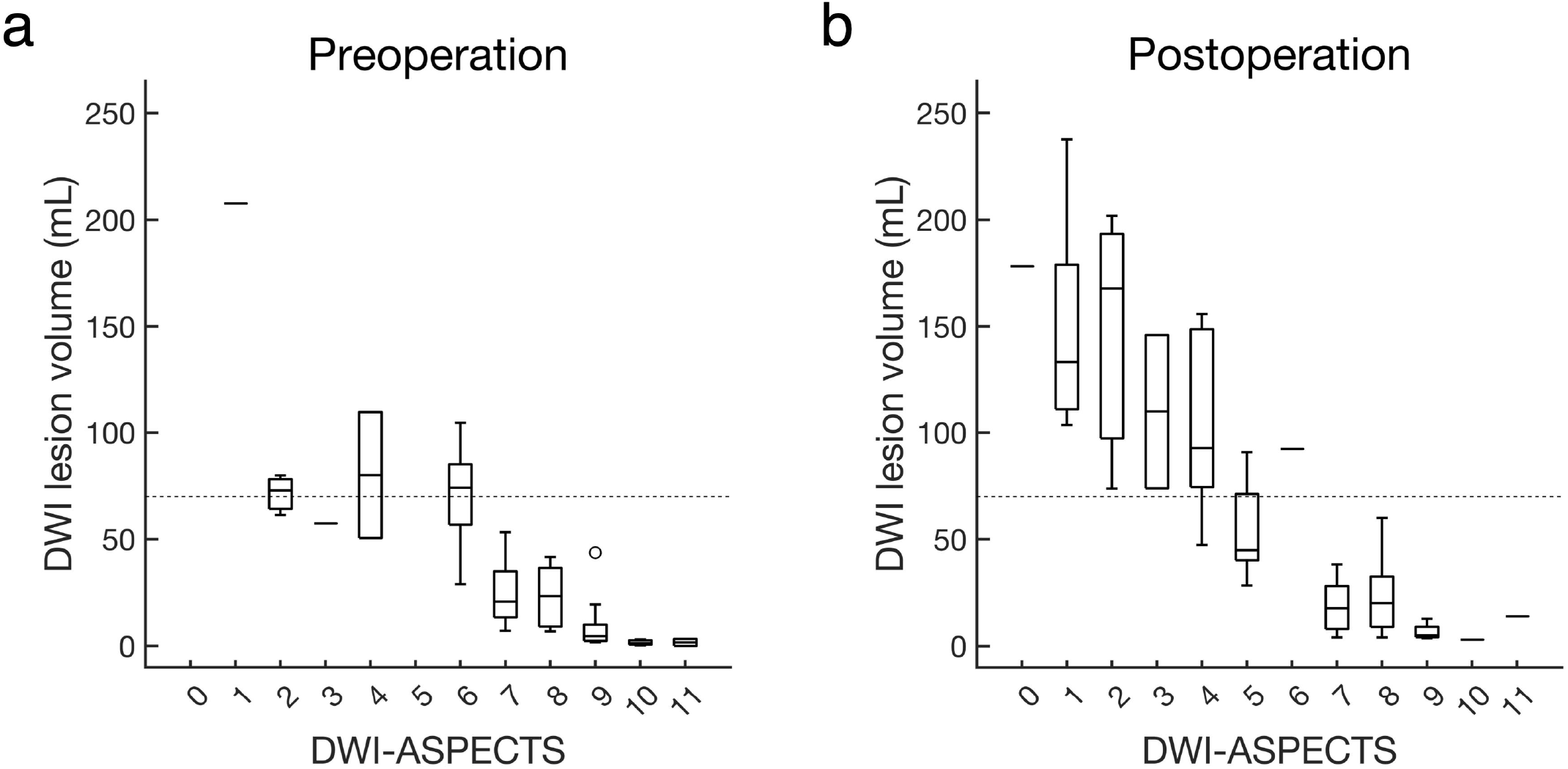
Box-and-whisker plot of DWI-ASPECTS and DWI lesion volume (Vol_DWI_) a. Values at preoperation. b. Values at postoperation. Broken horizontal lines indicate 70 mL of Vol_DWI_. Boxes indicate interquartile range; whiskers, extreme values; horizontal lines in each box, median; and a circle, outlier. DWI indicates diffusion-weighted imaging; and ASPECTS, Alberta Stroke Program Early Computed Tomography Score.

Next, we evaluated the differences between preoperative and postoperative values. In the good outcome group, the difference obtained from the preoperative minus the postoperative DWI-ASPECTS and c-ASPECTS was significantly lower than that in the not-good outcome group (*p* = 0.011 and 0.013, respectively: Wilcoxon rank-sum test). Moreover, in no changing groups, both DWI-ASPECTS and c-ASPECTS showed a significantly higher rate of a good outcome than worsening groups (*p* = 0.017 and 0.016, respectively: chi-squared test). The difference from the postoperative Vol_DWI_ minus the preoperative Vol_DWI_ in the good outcome group was significantly smaller than that in the not-good outcome group (*p* = 0.00086: Wilcoxon rank-sum test). Figure 2 indicates that the good outcome group showed no worsening of DWI-ASPECTS, c-ASPECTS, and Vol_DWI_ from preoperation to postoperation. Although the results of c-ASPECTS were similar to those of DWI-ASPECTS, results obtained from sc-ASPECTS showed no significance in preoperation, postoperation, and the difference. Additionally, we assessed the statistical difference between the no-changing group and the worsening group of c-ASPECTS (Supplementary Table 1). In the worsening group of c-ASPECTS, the baseline NIHSS and the retrieval attempts were significantly high (*p* = 0.032 and 0.045, respectively, Wilcoxon rank-sum test).

**Fig. 2.**
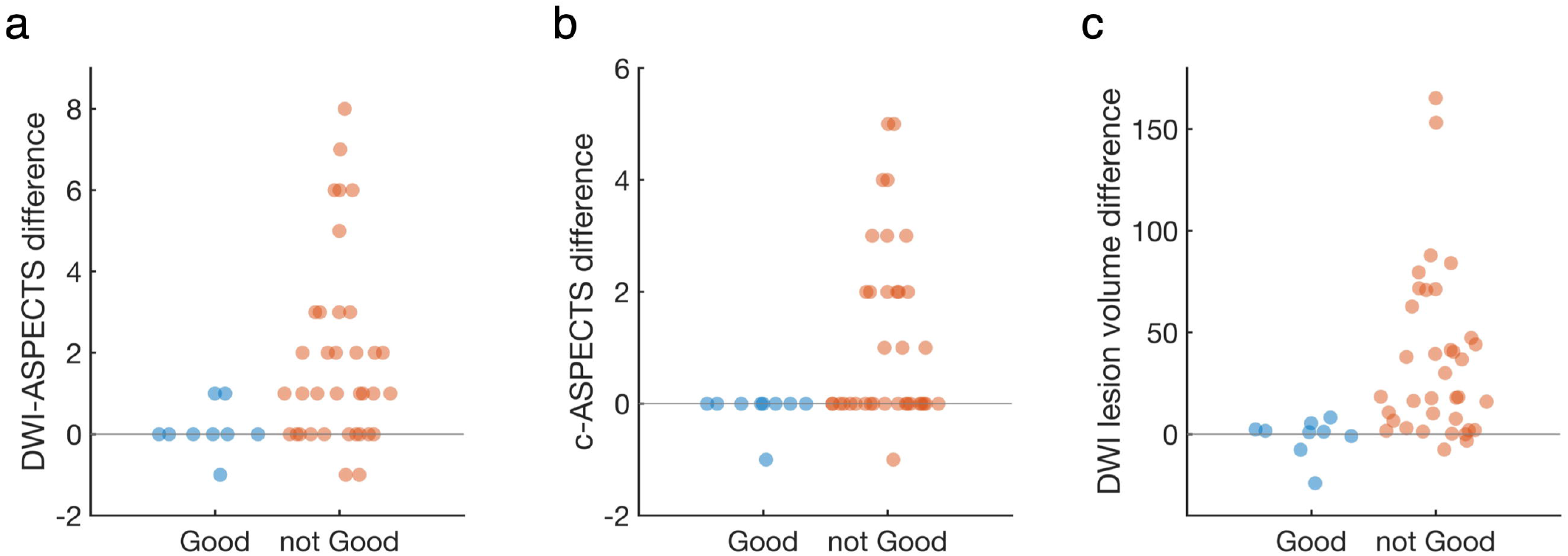
Beeswarm plot between the good and not-good groups The distribution patterns of differences between preoperation and postoperative values are shown. Differences obtained from preoperation minus postoperative values were used in DWI-ASPECTS (a), and c-ASPECTS (b). Differences obtained from postoperative minus preoperative values were used in DWI lesion volumes (c). ASPECTS indicates Alberta Stroke Program Early Computed Tomography Score; c-ASPECTS, cortical ASPECTS; and DWI, diffusion-weighted imaging.

### Factors for a good outcome

Table 3 indicates that NIHSS and postoperative DWI-ASPECTS were independent factors for a good outcome (OR = 0.69; 95% CI, 0.48 – 1.00; *p* = 0.046, OR = 2.27; 95% CI, 1.02 – 5.04; *p* = 0.039, respectively), as revealed by multivariate logistic regression analysis.

**Table. 3.** Multivariate logistic regression analysis for predictors of good outcomes (mRS 0 to 2).

| **Characteristics** | **OR (95% CI)** | ***p*-value** |
| --- | --- | --- |
| Age (years) | 0.91 (0.81 – 1.01) | 0.077 |
| NIHSS | 0.69 (0.48 – 1.00) | *0.046 |
| Postoperative DWI-ASPECTS | 2.27 (1.02 – 5.04) | *0.039 |

Statistically significant *p* values are flagged with an asterisk.
ASPECTS indicates Alberta Stroke Program Early Computed Tomography Score; CI, confidence interval; DWI, diffusion-weighted imaging; NIHSS, National Institutes of Health Stroke Scale; and OR, odds ratio

### ROC analysis

The COV of age and NIHSS by ROC curve analysis for predicting good outcomes was 75 years old (area under the curve (AUC) 0.87; sensitivity 0.72, specificity 0.89) and 13 (AUC 0.92; sensitivity 0.92, specificity 0.78), respectively (Figure 3a). The ROC curves obtained from DWI-ASPECTS and c-ASPECTS showed similarities. The COV obtained from the differences of DWI-ASPECTS and c-ASPECTS were 0 (AUC 0.77; sensitivity 0.67, specificity 0.78) and 0 (AUC 0.74; sensitivity 0.44, specificity 1.00), respectively (Figure 3b and 3c). In Vol_DWI_, ROC curves showed high performance; the COV of postoperation and difference was 20 mL (AUC 0.83; sensitivity 0.78, specificity 0.78), and 8 mL (AUC 0.88; sensitivity 0.69, specificity 1.00), respectively (Figure 3d).

**Fig. 3.**
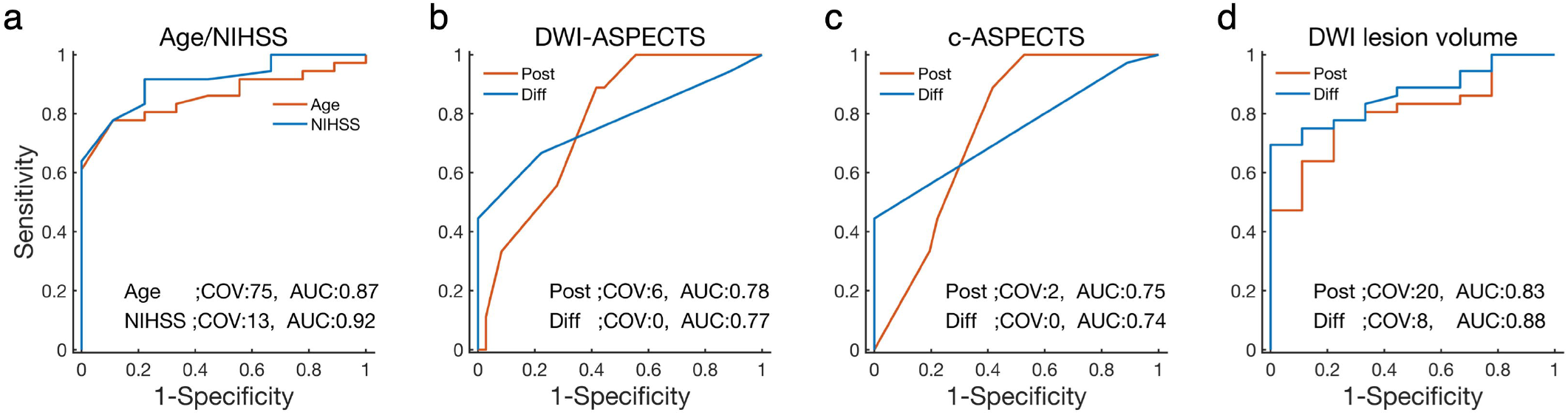
Receiver operating characteristic (ROC) curves ROC curves for predicting good outcomes (mRS: 0 or 2) are indicated and calculated from age and NIHSS (a), DWI-ASPECTS (b), c-ASPECTS (c), and Vol_DWI_ (d). Cut-off values (COV) and area under the curve (AUC) are shown. ASPECTS indicates Alberta Stroke Program Early Computed Tomography Score; c-ASPECTS, cortical ASPECTS; Diff, differences; DWI, diffusion-weighted imaging; mRS, modified Rankin Scale; NIHSS, National Institutes of Health Stroke Scale; Post, postoperation; and Vol_DWI_, DWI lesion volume.

## Discussion

We hypothesized that an assessment of changing DWI-ASPECTS from preoperation to postoperation would enable us to reveal a novel insight for predicting the outcome of AIS after MT. We could show that the good outcome group showed statistically no-changing DWI-ASPECTS and c-ASPECTS from before to after MT. Postoperative DWI-ASPECTS and c-ASPECTS and their differences showed good performance for predicting good outcomes through ROC analyses. The cut-off values for predicting good outcomes were 75 years old for age, 13 for NIHSS, and 0 for differences of DWI-ASPECTS and c-ASPECTS. Logistic regression analyses showed that postoperative DWI-ASPECTS and NIHSS were independent predictive factors for a good outcome.

In our institute, the AHA/ASA Guideline ^13^ was referred for selecting AIS patients to provide proper treatment. The guideline recommends MT to AIS because of anterior circulation LVO patients who show (1) pre-stroke mRS score of 0 to 1; (2) age ≥18 years; (3) NIHSS score of ≥6; and (4) ASPECTS of ≥6. However, a low score of DWI-ASPECTS before MT was associated with poor outcome ^14^ and a high score of that associated with good outcome ^20^, and the ESCAPE and REVASCAT trial excluded AIS patients with ASPECTS <6–7 or DWI-ASPECTS <6 ^5,8^. Thus, MT is not commonly recommended for large-volume AIS, and our results showed that DWI-ASPECTS <6 indicated zero percent of a good outcome. In this study, we set the inclusion criteria using preoperative DWI-ASPECTS. Our results showed no significant difference in preoperative DWI-ASPECTS between the good and not-good groups and that the median values of preoperative DWI-ASPECTS were over 6 scores. Our inclusion criteria that excluded the large volume AIS influenced these results, representing a selection bias.

Although preoperative DWI-ASPECTS has frequently been assessed as a factor for predicting outcomes after reperfusion therapy for AIS ^1,3,12,16,18,19,25-27^, studies investigating a relationship between the change in DWI-ASPECTS and reperfusion therapy for AIS are rare. The ischemic brain lesion extension on 24 hours was reported to be associated with the outcome of LVO-AIS ^16^. Our ROC analyses could add a novel insight that no-changing of DWI-ASPECTS from before to after MT distinguished good outcomes from the not-good outcomes. Moreover, we showed that the performance of the ROC curves in DWI-ASPECTS was similar to that in c-ASPECTS and that no change in c-ASPECTS was associated with a good outcome. A previous study showed a strong correlation between high c-ASPECTS and good outcomes in LVO-AIS, such as ASPECTS < 6 ^23^. Therefore, while this study indicated that postoperative DWI-ASPECTS and c-ASPECTS were associated with outcomes, we inferred that a true factor influencing the outcome after MT was c-ASPECTS. We propose that if physicians experience a worsening of c-ASPECTS after MT, they should be careful of poor outcomes.

While accuracy for identifying AIS was higher in DWI than in computed tomography (CT) ^28^, initial CT alone was reported to be non-inferior to initial CT plus additional MRI concerning outcomes ^29^. Moreover, since we showed that postoperative DWI-ASPECTS could distinguish good outcomes from not-good outcomes, we think an MRI examination before MT could be skipped to shorten the time from examination to treatment.

Recently, several studies demonstrated that MT was effective for even large-volume ischemic strokes, such as preoperative ASPECTS ≤5 ^23^/DWI-ASPECTS ≤ 5 ^25,30^. However, in large ischemic core stroke, older age ^25^ and large Vol_DWI_ ^25^ was associated with poor outcomes, and successful recanalization ^25,30-32^ was associated with a good outcome. Moreover, even at ASPECTS <6, high c-ASPECTS were reported to be a positive factor for good outcomes ^23^. Therefore, in our institute, even though patients showed large ischemic volumes such as DWI-ASPECT <6, we treated them by MT to realize successful recanalization. However, while we could achieve approximately 80% successful recanalization in this study, no patients showed good outcomes in preoperative DWI-ASPECTS <6. According to our results, the age of the not-good outcome group was significantly higher than that of the good outcome group. Patients aged ≥80 years after MT had worse functional outcomes and showed a lower percentage of successful recanalization ^33^. However, studies between 2017 and 2019 indicated better outcomes even in older patients ^33^, and the efficacy of MT for patients aged ≥80 years over standard care was demonstrated ^15,34^. A small infarct core and low NIHSS were associated with good outcomes in patients over 80 years of age ^20^. Although the efficacy of MT for AIS patients with low NIHSS scores is controversial, NIHSS was reported as a factor associated with outcomes after MT ^16^. A low NIHSS score was related to successful recanalization ^35^. We also showed that NIHSS was an independent factor for predicting good outcomes by the multivariate logistic regression analysis. Another factor related to the outcome after MT was the first-pass effect of MT, which is related to a favorable outcome ^36^. A previous study indicated that the first three retrieval attempts were associated with improved clinical outcomes ^37^. Our study could achieve less than three retrieval attempts in both the good and not-good outcome groups.

We demonstrated a strong negative correlation between DWI-ASPECTS and Vol_DWI._ All patients with DWI-ASPECTS ≥7, both preoperation and postoperation, had Vol_DWI_ <70 mL, in concordance with a previous study ^1^. Additionally, ASPECTS correlated with ischemic core volume ^11^, and Vol_DWI_ correlated strongly with final infarct volume ^28^. Therefore, DWI-ASPECTS and Vol_DWI_ are well-known factors related to the outcome of reperfusion therapy ^3,19,27^. We added new results that both preoperative and postoperative DWI-ASPECTS ≥7 may be reliable surrogates of Vol_DWI_ <70 mL.

Regarding sites of occlusion arteries, the M2 segment of MCA was assessed by MR CLEAN ^38^, ESCAPE ^5^, EXTEND-IA ^6^, and SWIFT PRIME ^7^ and was not assessed by REVASCAT ^8^, DAWN ^9^, and DEFUSE3 ^10^. Although the efficacy of MT for the M2 segment has been unclear ^15^, some studies showed the effectiveness of MT for LVO of the M2 segment ^39-42^. AIS attributed to the M2 segment of the MCA is known as medium vessel occlusion (MeVO) ^43^. While randomized studies have proven the safety and efficacy of MT for LVO ^5-8,38^, MT for MeVO has had no high-level evidence. However, since MeVO was observed in 25%–40% AIS ^44^ and showed substantial morbidity and the safety and efficacy of MT for MeVO have been presented ^45,46^, MT is increasingly performed for AIS-MeVO ^47^ in actual clinical situations. Therefore, our study included M2 occlusion, and there was no significant difference in arterial occlusion site, while a low percentage of the good outcome was observed in the M2 occlusion.

This study has some limitations. First, a small amount of our data was retrospectively collected from a single center. No control group treated with conventional medical care was enrolled, and the study was nonrandomized. Second, we focused on only AIS because of the anterior circulation occlusion, including CCA, ICA, M1, and M2 segments. Therefore, our findings could not be applied to AIS because of the posterior circulation occlusion. Third, we defined MRI obtained within 21 postoperation days as postoperative MRI. The number 21 was decided by our empirical experience, and it remains unclear how long after MT was useful for predicting outcomes. Finally, we did not measure the volume of the ischemic penumbra area before MT. While we revealed that the worsening of the c-ASPECTS group showed a statistically high score of NIHSS, it is possible that the worsening of c-ASPECTS is associated with the volume of the ischemic penumbra area because the volume of DWI-MRI and perfusion-weighted MRI correlated positively with NIHSS score ^48^.

In conclusion, an assessment of changes in DWI-ASPECTS from before to after MT revealed that the good outcome group showed no change in DWI-ASPECTS and c-ASPECTS. The independent factors for predicting a good outcome after MT were low NIHSS and high postoperative DWI-ASPECTS. The cut-off values for a good outcome were 75 years old, NIHSS of 13, and a difference in DWI-ASPECTS of 0.

## Non-standard Abbreviations and Acronyms

ASPECTS: Alberta Stroke Program Early Computed Tomography Score
CCA: common carotid artery
DWI: diffusion-weighted imaging
ICA: internal carotid artery
IQR: interquartile range
MCA: middle cerebral artery
mRS: modified Rankin Scale
mTICI: modified Thrombolysis in Cerebral Infarction
NIHSS: National Institutes of Health Stroke Scale
SAH: subarachnoid hemorrhage
tPA: tissue plasminogen activator

## Acknowledgments

We want to thank Dr. Masayuki Hirata at the Department of Neurological Diagnosis and Restoration at the Graduate School of Medicine, Osaka University, for his technical support and Dr. Haruhiko Kishima at the Department of Neurosurgery at Osaka University for his comment.

## Author Contributions

HH conceived this study, collected the data, created the MATLAB program, analyzed the data, created all figures, and was primarily responsible for writing the manuscript. HH, TM, and HN discussed the interpretation of the results. TM supervised the study. All authors reviewed the manuscript.

## Supplemental Material

Table S1

