## Supplementary Table 1 for "No changes in diffusion-weighted imaging-Alberta Stroke Program Early Computed Tomography score from before to after mechanical thrombectomy for anterior circulation occlusion are associated with good outcomes"

**Supplementary Table. 1** Descriptive statistics regarding baseline, procedural, and outcome parameters related to c-ASPECTS changes.

| **Characteristics** | **Patients n (%)** | **No changes (% or IQR)** | **Worsening (% or IQR)** | ***p*-value** |
| --- | --- | --- | --- | --- |
| **Sex** |  |  |  |  |
| Male | 22 (49) | 14 (64) | 8 (36) | 0.91 |
| Female | 21 (47) | 13 (62) | 8 (38) |  |
| **Age (years)** |  | 76 (61 – 86) | 85 (69 – 88) | 0.57 |
| **NIHSS** |  | 17 (12 – 21) | 20 (18 – 22) | *0.032 |
| **Premorbid mRS score** |  | 2 (0 – 4) | 0 (0 – 1) | 0.055 |
| **Laterality** |  |  |  |  |
| Left | 24 (53) | 15 (62.5) | 9 (37.5) | 0.97 |
| Right | 19 (42) | 12 (63) | 7 (37) |  |
| **Arterial occlusion site** |  |  |  |  |
| CCA + ICA + MCA-M1 | 30 (67) | 21 (70) | 9 (30) | 0.14 |
| MCA-M2 | 13 (29) | 6 (46) | 7 (54) |  |
| **Retrieval attempts** |  | 1 (1 – 2) | 2(2 – 3) | *0.045 |
| **Onset to reperfusion time (hours)** |  | 6 (4 – 10) | 5 (4 – 8) | 0.31 |
| **mTICI grade** |  |  |  |  |
| 2b + 3 | 35 (78) | 22 (63) | 13 (37) | 0.99 |
| Others | 8 (18) | 5 (62.5) | 3 (37.5) |  |
| **tPA therapy** |  |  |  |  |
| Yes | 17 (38) | 8 (47) | 9 (53) | 0.084 |
| No | 26 (58) | 19 (73) | 7 (27) |  |
| **Preoperative DWI-ASPECTS** |  |  |  |  |
| ≥6 | 36 (80) | 21 (58) | 15 (42) | 0.17 |
| <6 | 7 (16) | 6 (86) | 1 (14) |  |
| **Hemorrhagic infarction** |  |  |  |  |
| Yes | 23 (51) | 13 (57) | 10 (43) | 0.36 |
| No | 20 (44) | 14 (70) | 6 (30) |  |
| **SAH** |  |  |  |  |
| Yes | 11 (24) | 6 (55) | 5 (45) | 0.51 |
| No | 32 (71) | 21 (66) | 11 (34) |  |

Patients (%) or median (IQR) are demonstrated for categorical data or continuous variables, respectively. *P* values were calculated using the chi-squared test for categorical variables and the Wilcoxon rank-sum test for continuous variables. Statistically significant *p* values are flagged with an asterisk.

ASPECTS indicates Alberta Stroke Program Early Computed Tomography Score; c-ASPECTS, cortical ASPECTS; CCA, common carotid artery; DWI, diffusion-weighted imaging; ICA, internal carotid artery; IQR, interquartile range; MCA-M1, middle cerebral artery-M1 segment; MCA-M2, middle cerebral artery-M2 segment; mRS, modified Rankin Scale; mTICI, modified Thrombolysis in Cerebral Infarction; NIHSS, National Institutes of Health Stroke Scale; SAH, subarachnoid hemorrhage; and tPA, tissue plasminogen activator.
